# HiFiMAP: High-resolution fast identity-by-descent mapping test

**DOI:** 10.64898/2026.05.06.26352570

**Authors:** Bohong Guo, Ardalan Naseri, Ziqian Xie, Chloé Sarnowski, Degui Zhi, Han Chen

## Abstract

Although traditional genome-wide association studies (GWAS) have identified numerous loci, they often ignore phased haplotype information. Identity-by-descent (IBD) mapping captures these extended haplotypic effects by modeling shared ancestral segments. However, standard statistical mapping of these segments scales poorly with biobank-sized cohorts and short IBD segments that capture older evolutionary events. To overcome this computational bottleneck, existing scalable IBD mapping frameworks aggregate shared segments into fixed sliding windows. While computationally efficient, this window-based approach generates association signals at a low resolution that often span hundreds of kilobases. To address this issue, here we present a novel High-resolution Fast IBD Mapping test (HiFiMAP) that takes snapshots of IBD segments at the single nucleotide polymorphism (SNP) level resolution. Simulation studies confirm that HiFiMAP maintains well-controlled type I error rates and exhibits superior statistical power for detecting rare variants and haplotype effects using short IBD segments. In a UK Biobank (UKB) benchmark (N=407,681), HiFiMAP mapped 640,899 SNPs at 1.92 CPU seconds per test, massively outperforming existing window-based methods (95.2 CPU seconds per test for 3,403 windows). Furthermore, applied to high-dimensional brain imaging phenotypes (N∼36,000), HiFiMAP identified five novel associations previously undetected by standard GWAS approaches, including key central nervous system regulators like *NR2F1* and *NSF/WNT3*. By refining large testing windows into highly specific genomic variants, HiFiMAP empowers biobank-scale, SNP-level resolution mapping to accurately pinpoint complex trait architectures.

## Introduction

Over the past two decades, genome-wide association studies (GWAS) have revolutionized our understanding of the genetic basis of complex human diseases and quantitative traits^1,2^. By leveraging biobank-scale cohorts like the UK Biobank (UKB)^3^, researchers have identified thousands of genetic loci associated with brain morphology and neurological disorders^4–6^. However, a significant portion of the heritability for these traits remains unexplained. Traditional GWAS approaches primarily focus on independent single-nucleotide polymorphisms (SNPs) and often ignore phased haplotype information^1,7^. While variant set tests like the cohort allelic sums test (CAST), the sequence kernel association test (SKAT), and others^8–14^ have been developed to aggregate rare variant signals from sequencing data, they typically evaluate these mutations as unphased entities within a region. Consequently, they are intrinsically limited in capturing the collective impact of extended haplotype effects.

Identity-by-descent (IBD) mapping offers a powerful, complementary alternative to traditional GWAS by leveraging the shared ancestral segments between individuals^15^. Because IBD segments represent long, contiguous haplotypes inherited from a common ancestor, they are naturally more robust against genotyping or sequencing errors and can capture the effects of untyped rare variants that occur on these shared backgrounds^15,16^. Previous studies have demonstrated that IBD mapping can reveal genetic associations and “hidden” heritability that standard GWAS fails to detect, particularly in the context of rare variations and haplotype effects^7,17^. Despite the development of highly efficient algorithms for IBD segment detection^16,18–21^, which have largely resolved the O(N^2^) complexity of IBD calling, the statistical mapping of these segments in biobank-scale cohorts remained computationally prohibitive due to the O(N^3^) time and O(N^2^) memory complexity of traditional variance component models^7,15,22,23^. To account for this main computational challenge, new IBD mapping approaches have recently been developed for biobank-scale cohorts^7,24^. These advancements empower efficient IBD mapping for complex traits across unprecedented sample sizes.

However, low resolution is a common limitation for existing IBD mapping methods. For example, FiMAP chunks the genome into fixed testing windows, such as 1 centimorgan (cM) segments^7^. While Cai and Browning narrowed this sliding-window approach down to 0.1 cM segments^24^, these window-based frameworks still yield a lower resolution compared to the single-SNP precision of traditional GWAS. often leaves researchers with broad genomic regions that are difficult to fine-map or interpret. Furthermore, the limitations of window-based IBD mapping are further amplified when researchers attempt to capture older coalescence events by lowering the IBD detection threshold,. the minimum length of a shared haplotype required by the IBD caller. Lowering the detection threshold from 3 cM used in FiMAP to 2 cM would exponentially increase the density of the IBD sharing matrix due to the inclusion of older, more prevalent haplotypes and background linkage disequilibrium (LD). This increased matrix density pushes existing window-based methods like FiMAP^7^ to their computational limits. The challenge of resolution and efficiency is further amplified when analyzing high-dimensional phenotypes, such as brain imaging data from the UKB^25–27^. Modern neuroimaging studies generate hundreds to thousands of imaging-derived phenotypes (IDPs) and unsupervised deep-learning-derived imaging phenotypes (UDIPs)^28–31^.

Mapping IBD associations for these high-dimensional complex brain traits demands a new level of computational efficiency to integrate genetic findings with neurobiological mechanisms.

To address these compounding challenges, we propose the High-resolution Fast IBD Mapping test (HiFiMAP). Instead of IBD windows of fixed lengths, HiFiMAP takes a snapshot of IBD sharing at each SNP and introduces a dynamic updating algorithm that achieves SNP level resolution while also greatly boosting the computational efficiency. Rather than redundantly calculating variance components for arbitrary fixed windows, HiFiMAP updates the IBD matrix exclusively at the genomic positions where IBD segments start or end. To establish the validity and enhanced statistical power of our approach, we applied HiFiMAP to six well-characterized anthropometric traits in a cohort of 407,681 White British individuals from the UKB which were previously analyzed using FiMAP. Using IBD segments with lengths of at least 3 cM, we demonstrated that HiFiMAP is highly consistent with the FiMAP results using the same IBD segments^7^. Building on this consistency, our further analysis using IBD segments with lengths of at least 2 cM produced much stronger association signals and identified novel loci missed by analysis using IBD segments with lengths of at least 3 cM and the GWAS. We also leveraged this highly scalable foundation and applied HiFiMAP to high-dimensional brain imaging data utilizing IBD segments with lengths of at least 1 cM. Specifically, we analyzed 128 T1/T2 UDIPs extracted from 36,241/35,111 UKB participants. By capturing older, ancient IBD segments, this high-resolution approach successfully uncovered multiple novel genome-wide significant associations linking regional brain morphology and pathways underlying severe neurodegenerative disorders that are missed by single-variant GWAS, thereby providing a more comprehensive view of the genetic drivers of brain morphology.

## Methods

### HiFiMAP

The original FiMAP methodology was based on a variance component model with finite-sample adjustment and computational approximation to enhance efficiency for large datasets^7^. Briefly, let *r* = *Y* − *Xβ̂* − *b̂* be the length *N* residual vector from a fitted linear mixed model under the null hypothesis of no local IBD associations, where *Y* represents the phenotype, *X* are the covariates, *β̂* and *b̂* are fixed effects and polygenic random effects estimated from this model, and *σ̂*^2^ is the estimated residual variance. A classical score-type variance component test for IBD association at locus *l* is given by 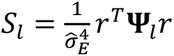, where **Ψ***_l_* is the local IBD matrix defined as the copies of the genome shared IBD at this genetic locus *l*. Combined with finite-sample adjustment and computational approximation, this model enables efficient hypothesis testing for local IBD associations while accounting for global relatedness and covariate effects^7^.

Here we illustrate the main methodological advances for efficient SNP-level IBD mapping using HiFiMAP. Building on FiMAP, the HiFiMAP method focuses on developing an efficient SNP-level IBD mapping test to investigate the genetic architecture of individual phenotypes that is complementary to GWAS. We compute the variance component test statistic *S*_*j*_ using **Ψ**_*j*_, the IBD matrix at SNP *j*. To improve computational efficiency, SNP-level IBD matrix is updated dynamically based on overlapping IBD segments:

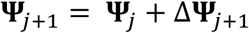

Where **Ψ**_*j*_ is the current IBD matrix at SNP *j*, Δ**Ψ**_*j*+1_ denotes the changes from **Ψ**_*j*_ to the IBD matrix **Ψ**_*j*+1_ for SNP *j* + 1. The IBD segments identified from the IBD callers are sorted by their start positions and end positions to ensure they are efficiently processed by this dynamic updating algorithm. The sorting prevents redundant recalculations of contributions from IBD segments that persist across consecutive positions. Therefore, at each test location (SNP position), this algorithm checks whether to add segments starting this position, and whether to subtract segments that end before this position. In this way, it ensures that the IBD matrix reflects the cumulative effect of overlapping segments at the specific genomic location, without complete recalculations. A representative example is illustrated in Fig 1, which shows the sequential update of the test statistics across three SNPs.

**Fig 1.**
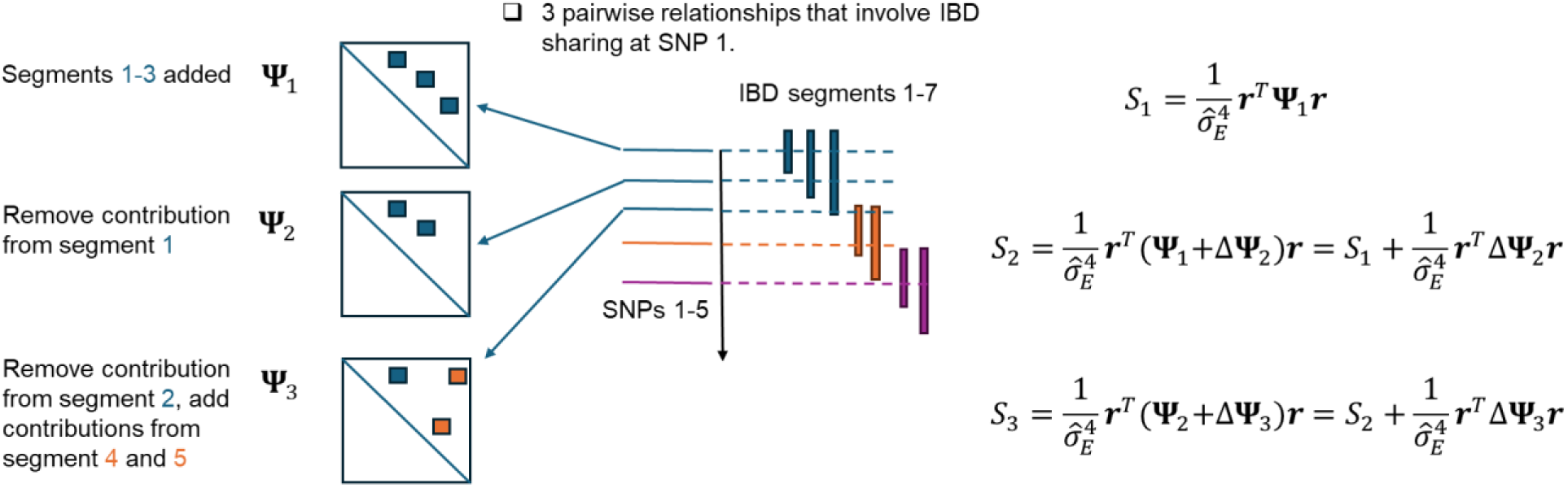
Illustration of the dynamic updating algorithm employed for efficient IBD mapping. The schematic demonstrates how the IBD sharing matrix **Ψ** and the test statistic ***S*** are updated incrementally as the analysis progresses along genomic markers (SNPs 1-5). Rather than recalculating the full matrix at every position, the algorithm efficiently modifies the previous statistic by adding or removing only the contributions of IBD segments that start or end at the current SNP (Δ**Ψ**).

To mitigate the severe I/O bottlenecks associated with repeatedly parsing massive, biobank-scale IBD segment files, we developed a specialized IBD reformatting tool. By pre-parsing and saving genomic checkpoints (local IBD matrices), this tool allows us to precisely partition chromosomes at these indexed sites. This structural chunking enables the framework to fully leverage distributed computing environments, facilitating the massive parallelization of IBD mapping across high-performance computing (HPC) clusters and significantly reducing overall runtimes.

### Simulations

To validate the type I error rates and statistical power of HiFiMAP approach, we conducted extensive simulations using synthetic datasets. We utilized coalescent simulations via msprime^32^ to generate whole-genome sequencing data containing 20,421,461 genetic variants for a cohort of 5,000 individuals. We modeled realistic genetic variation and population haplotype structures using the HapMap phase II GRCh37 recombination map and a mutation rate of 1.38 × 10^−8^. From the simulated haplotypes, we utilized hap-IBD to infer IBD segments at three different minimum length thresholds: ≥1 cM, ≥2 cM, and ≥3 cM. Before proceeding to association testing, we evaluated the accuracy of these inferred segments against the msprime ground-truth utilizing an established open-source IBD benchmarking tool^33^. Accuracy and length accuracy were recorded for IBD segments with 1 cM, 2 cM and 3 cM thresholds. The accuracy is defined as the number of covered IBD segments divided by the total number of reported IBD segments, the length accuracy is a more fine-grained measure for reported IBD segments, which accounts for the portions of falsely reported segments along a true IBD segment^33^.

To assess type I error rates under the null hypothesis of no local IBD random effects, a continuous phenotype *Y*_*i*_ for individual *i* was simulated as *Y*_*i*_ = 0.1*age*_*i*_ + *sex*_*i*_ + *b*_*i*_ + *ɛ*_*i*_, where the vector form *b* followed a multivariate normal distribution with mean 0 and covariance matrix ɸ from the kinship matrix of the simulated cohort, and *ɛ*_*i*_ followed a standard normal distribution. Age is simulated from a normal distribution with mean 50 and standard deviation 5, and sex from a Bernoulli distribution with probability 0.5.

Crucially, because the test statistic remains identical across intervals where the IBD matrix is constant, hypothesis testing was restricted to SNPs where the local IBD sharing changed. We simulated 200 phenotype replicates to obtain a total of 1,043,096,000, 624,589,400 and 440,461,200 p-values respectively for 1 cM, 2 cM and 3 cM analysis under the null hypothesis of no IBD random effects.

We then benchmarked the statistical power of GWAS and HiFiMAP across the three IBD length thresholds using two distinct causal scenarios: rare variant effects and haplotype effects. Specifically, for rare variant effects, in each simulation replicate we randomly selected a 1 cM window with at least 8 ultra-rare variants (MAF < 0.0005); we then randomly assigned 8 ultra-rare variants in this window as causal variants and simulate a continuous phenotype *Y*_*i*_ for individual *i* as 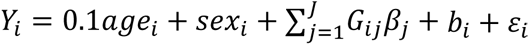, where *age*_*i*_, *sex*_*i*_, *b*_*i*_ and *ɛ*_*i*_ were simulated using the same parameter settings as in the type I error simulations, *G*_*ij*_ represents the genotype of individual *i* for the causal variant *j*, and the causal variants effect is *β*_*j*_ 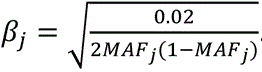. Empirical power was estimated as the proportion of any HiFiMAP p values for those ultra-rare causal variants being less than the simulated IBD mapping genome-wide significance level^34^ of 2.32 × 10^−6^, 3.54 × 10^−6^ and 5.27 × 10^−6^ for 1 cM, 2 cM and 3 cM analysis respectively, over 1000 replicates. For GWAS running on the same replicates, we took the phenotypic residuals estimated from the HiFiMAP null model and performed standard single-variant GWAS for all common variants (MAF>0.01) within the 1 cM causal window. We added a 10 kb flanking region upstream and downstream of each of the 8 simulated causal variants within the 1 cM window. Minimum p-values were taken for each 20kb localized window to represent the significance of each causal variant. Empirical GWAS power was then calculated as the proportion of p-values for those causal variants achieved the standard genome-wide significance threshold of 5 × 10^−8^.

Additionally, we simulated haplotype effects. We estimated all haplotypes with length ≥ 1 cM shared by at least 30 individuals using the phased genotypes of the 5,000 simulated individuals and the PBWT-block algorithm^35,36^. In each simulation replicate, we randomly sampled one of these haplotypes and assigned it as causal. And then simulate the continuous phenotype *Y*_*i*_ for individual *i* as: *Y*_*i*_ = 0.1*age*_*i*_ + *sex*_*i*_ + *H*_*i*_*γ* + *b*_*i*_ + *ɛ*_*i*_, same simulation setting was applied for *age*_*i*_, *sex*_*i*_, *b*_*i*_ and *ɛ*_*i*_, and *H*_*i*_ (0/1/2) is the number of causal haplotypes carried by individual *i*. The causal haplotype effect was assigned as 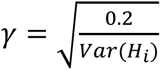. Empirical power was estimated as the proportion of replicates where at least one variant within the simulated causal haplotype region being less than the simulated IBD mapping genome-wide significance level^34^ of 2.32 × 10^−6^, 3.54 × 10^−6^ and 5.27 × 10^−6^ for 1 cM, 2 cM and 3 cM analysis using hap-IBD segment calls respectively, over 1000 replicates. We performed GWAS on the same replicates similarly as the rare-variants scenario using phenotypic residuals, empirical power was estimated as the proportion of replicates where at least one variant within the simulated causal haplotype region was significant at the 5 × 10^−8^ threshold.

### Real data analysis

For our empirical analyses, we utilized the UKB version 2 genotyping array data (ukb_hap_chr1-22_v2.bgen)^3^. We applied the comprehensive quality control (QC) procedures established by the UKB genetics team, restricting our cohort to individuals of White British ancestry to minimize the confounding effects of population structure. Our analysis was approved by the UTHealth Houston committee for the protection of human subjects. UKB has secured informed consent from the participants in the use of their data for approved research projects. To extract the haplotype phase information necessary for our method, IBD segments were called across the autosomes using the highly efficient software programs hap-ibd^16^ and RaPID^18^. The segment calling was guided by the standard GRCh37 genetic recombination map. To ensure direct comparability with the established FiMAP baseline, we extracted the same six well-characterized anthropometric traits from the UKB: standing height, body mass index (BMI), weight, waist circumference, hip circumference, and sitting height. We adopted the same quality control procedures and covariate adjustment as the original FiMAP study, adjusting for age, age^2^, sex, their interactions, and top 10 ancestral principal components. This rigorous alignment resulted a finalized cohort of White British individuals, including sample sizes of 407,872 for Waist circumference, 407,827 for Hip circumference, 407,681 for Standing height, 407,323 for Sitting height, 407,255 for Body mass index, and 407,400 for Body weight. This large-scale dataset served as the foundation for validating the SNP level resolution of HiFiMAP against known 3 cM association peaks (3 cM analysis), as well as testing its empirical performance at the 2 cM threshold (2 cM analysis).

To evaluate the performance of HiFiMAP on high-dimensional brain imaging data with 1 cM IBD segment threshold, we utilized the UDIPs from Patel et al^28^. These 128 T1/T2 UDIPs were generated from structural brain magnetic resonance imaging (MRI) scans. We had T1/T2 UDIPs data for a subset of 36,241/35,111 White British individuals from UKB. Prior to genetic association testing, each UDIP was adjusted for standard covariates (including age, sex, and the first 10 ancestral principal components). The resulting phenotypic residuals were then used for subsequent IBD mapping. We reported the minimum p-value (minP) across the 128 UDIPs for each SNP. Additionally, we sought to determine if the HiFiMAP signals remained significant after regressing out the effects of the top independent GWAS common variants located within the respective genomic regions. To achieve this, we conducted a rigorous conditional analysis. We first identified all GWAS variants exceeding the unadjusted genome-wide significance threshold (*p* < 5 × 10^−8^) within the ± 1 megabase (Mb) region of each HiFiMAP peak and pruned them for LD (*r*^2^ > 0.5, physical distance < 100 kb) to establish a set of independent associations. We then ran a null model for each UDIP dimension that was driving the minP signal, explicitly including the corresponding GWAS SNPs as additional covariates. Lastly, we performed the HiFiMAP test using the scaled residuals from these null models for each of the above UDIP dimensions.

### Multiple testing adjustment for IBD mapping

In traditional GWAS, the genome-wide significance threshold is conventionally set to 5 × 10^−8^ to account for the multiple testing burden of approximately one million independent common variants^1,37^. However, applying this standard threshold to IBD mapping is overly conservative^24^. Unlike LD, which decays rapidly across the genome, IBD segments represent long, contiguous blocks of shared ancestry that decay much more slowly. This biological property results in a highly correlated test statistic structure across adjacent genomic regions, drastically reducing the number of effective independent tests. For the empirical analysis of the UKB anthropometric traits, we adopted the genome-wide significance thresholds specifically established for IBD segments by Browning et al^34^. For the White British cohort consisting of approximately 410,000 individuals, the threshold was recommended as 2.13 × 10^−6^ for 2 cM segments and 2.70 × 10^−6^ for 3 cM segments. For the simulation studies utilizing a synthetic cohort of 5,000 individuals, we followed the theoretical procedures established by Browning et al.^34,38^ to estimate the appropriate, properly calibrated genome-wide significance levels for the 1 cM, 2 cM, and 3 cM simulation scenarios.

## Results

### Simulation study

The QQ plot for the null simulation is shown in Fig 2, HiFiMAP demonstrated well-calibrated test statistics across all three thresholds (*λ*_*GC*_ ranging from 1.012 to 1.025). The empirical type I error rates were 2.71 × 10^−6^, 3.99 × 10^−6^ and 3.51 × 10^−6^ at significance levels of 2.32 × 10^−6^, 3.54 × 10^−6^ and 5.27 × 10^−6^ (Supplementary Table 1) for 1 cM, 2 cM and 3 cM analysis respectively, indicating proper type I error controls.

**Fig 2.**
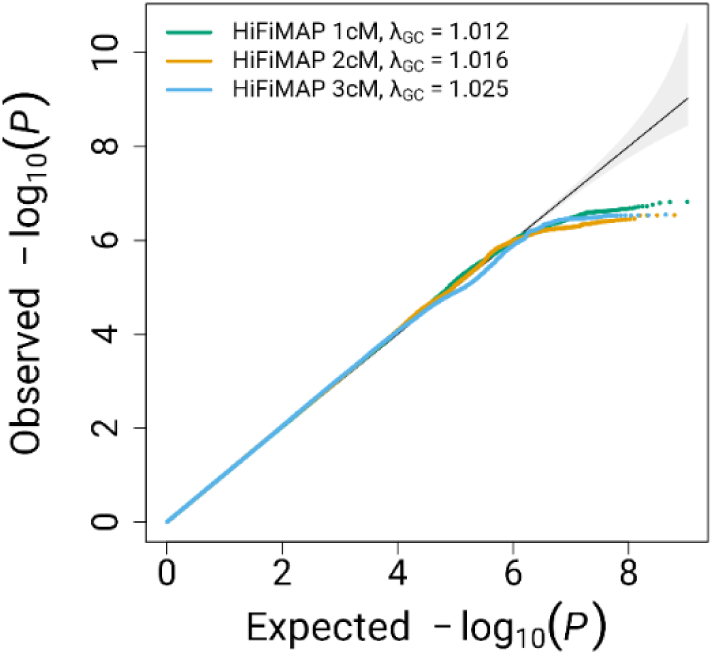
HiFiMAP 1 cM (green) and HiFiMAP 2 cM (orange) and HiFiMAP 3 cM (blue) QQ plot under the null hypothesis of no genetic association with a sample of 5000 individuals.

The power comparison for standard GWAS and HiFiMAP across different IBD length thresholds for rare variants-based effects and haplotype-based effects are shown in Fig 3. On one hand, the results of these power simulations highlight the advantage of utilizing shorter segments in IBD association mapping, particularly for detecting haplotype-driven associations. Under the rare variant scenario, power remained highly comparable and stable across all three thresholds (88.1% for 1 cM, 86.7% for 2 cM, and 86.2% for 3 cM), indicating that the choice of length threshold does not severely impact the detection of highly localized ultra-rare variant burdens. While for GWAS, the power is 58.7%, since the causal ultra-rare variants are routinely filtered out during standard GWAS procedures. Therefore, single-variant tests must rely on local LD with common tag SNPs to capture the association signal.

**Fig 3.**
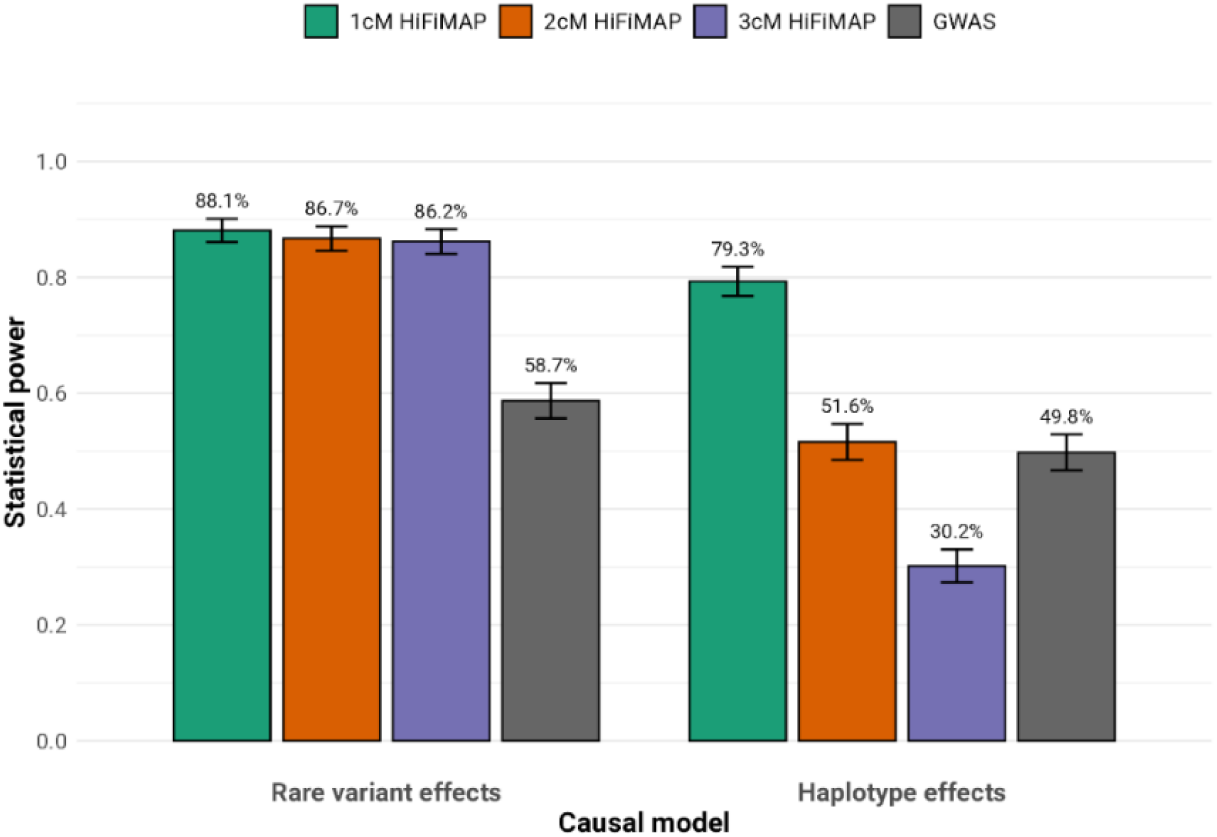
Statistical power of GWAS and HiFiMAP utilizing 1 cM, 2 cM, and 3 cM IBD length thresholds under two simulated causal scenarios. The rare variant model (left) simulated 8 causal ultra-rare variants (MAF < 0.0005) within a 1 cM window. The haplotype model (right) simulated a ≥1 cM causal haplotype present in at least 30 carriers.

Conversely, under the haplotype effect scenario, the minimum IBD length threshold played a highly influential role. Statistical power dropped steeply as the stringency increased, falling from 79.3% at 1 cM down to 51.6% at 2 cM, and reaching a low of 30.2% at 3 cM. The standard GWAS achieved 49.8% empirical power, comparable to 2 cM HiFiMAP but exceeded 3 cM HiFiMAP. This is probably because the broader testing windows (the entire causal haplotype region) for GWAS make it easier for common tag SNPs to partially capture the haplotype effects. Furthermore, the substantial reduction in power at longer length cutoffs for HiFiMAP occurs because the stricter 2 cM and 3 cM cutoffs fail to capture a large proportion of the shorter IBD segments shared among carriers of the causal haplotype. By lowering the threshold to 1 cM, HiFiMAP successfully captures a much higher density and volume of local IBD sharing in the simulated cohort. This higher abundance of IBD segments at the causal locus provides the necessary genetic signals to achieve substantial gain in statistical power. However, the 1 cM threshold yielded lower segment accuracy (accuracy 0.76, length accuracy 0.72) compared to the more stringent 2 cM (accuracy 0.99, length accuracy 0.96) and 3 cM (accuracy 0.99, length accuracy 0.97) thresholds (Supplementary Table 2). This indicated that slightly lower IBD segments accuracy at 1 cM does not adversely affect the detection of ultra-rare variants or haplotype effects. The increased density of local IBD sharing captured at the 1 cM threshold might outweigh the noise introduced by the lower calling accuracy during downstream association testing. By incorporating these 1 cM segments in this small cohort, HiFiMAP unlocks a massive power gain for detecting shorter shared haplotypes, firmly justifying the transition to high-resolution, lower-threshold IBD mapping.

### Application to UKB anthropometric traits

Compared to our previous tool FiMAP which gives IBD mapping results based on 1 cM chunks over the genome and shows complementary association evidence from GWAS, HiFiMAP tests each IBD segment change point in a study sample and can reach single SNP resolution in biobank scale data. Fig 4 shows that HiFiMAP single SNP resolution results had similar patterns with previous FiMAP 3 cM resolution results on standing height (N = 407,681) and sitting height (N = 407,323). It took HiFiMAP 344 CPU hours to compute 640,899 p-values, compared to 90 CPU hours for 3,403 p-values using FiMAP.

**Fig 4.**
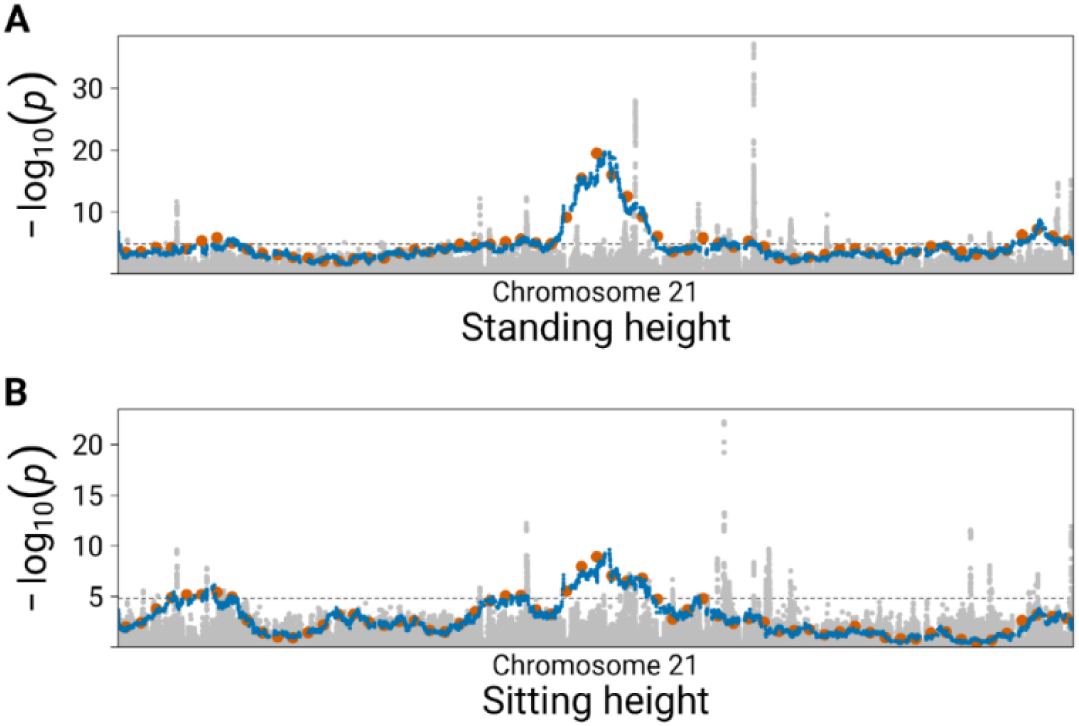
GWAS (grey), FiMAP (orange) and HiFiMAP (blue) p-values for standing and sitting height on chromosome 21. (A) Standing height; (B) Sitting height. IBD segments called by RaPID with a length of at least 3 cM were used in the FiMAP (1 cM resolution) and HiFiMAP (single SNP resolution) analyses. The dashed line represents the significance threshold from FiMAP: 0.05/3,403 = 1.47 × 10^−5^.

To further evaluate the impact of IBD segment length thresholds on the strength of the association signals, we compared HiFiMAP results using 2 cM and 3 cM cutoffs against standard GWAS baseline results. Figure 5 illustrates these comparisons across four distinct loci and anthropometric traits. Panel A highlights the well-known *FTO* locus for body mass index on chromosome 16, where the 2 cM HiFiMAP analysis yielded a highly significant association signal that substantially exceeded the 3 cM HiFiMAP analysis. This trend of amplified statistical significance for the 2 cM analysis was consistent across other prominent regions, including *PAPPA2* locus for standing height (panel B), *UQCC1/GDF5* locus for waist circumference (panel C), and *ACAN* locus for sitting height (panel D). Notably, at the *PAPPA2* locus, 2 cM analysis produced much stronger signals than 3 cM analysis and GWAS. And at the *UQCC1/GDF5* locus , the 2 cM HiFiMAP analysis clearly surpassed the established significance threshold (2.13 × 10^−6^), whereas both the 3 cM analysis and standard GWAS yielded much weaker, non-significant association signals. Overall, these regional plots demonstrate that utilizing a 2 cM threshold can capture critical associations that are attenuated at longer IBD segment length cutoffs (3 cM analysis) and undetected by traditional single-variant approach (GWAS).

**Fig 5.**
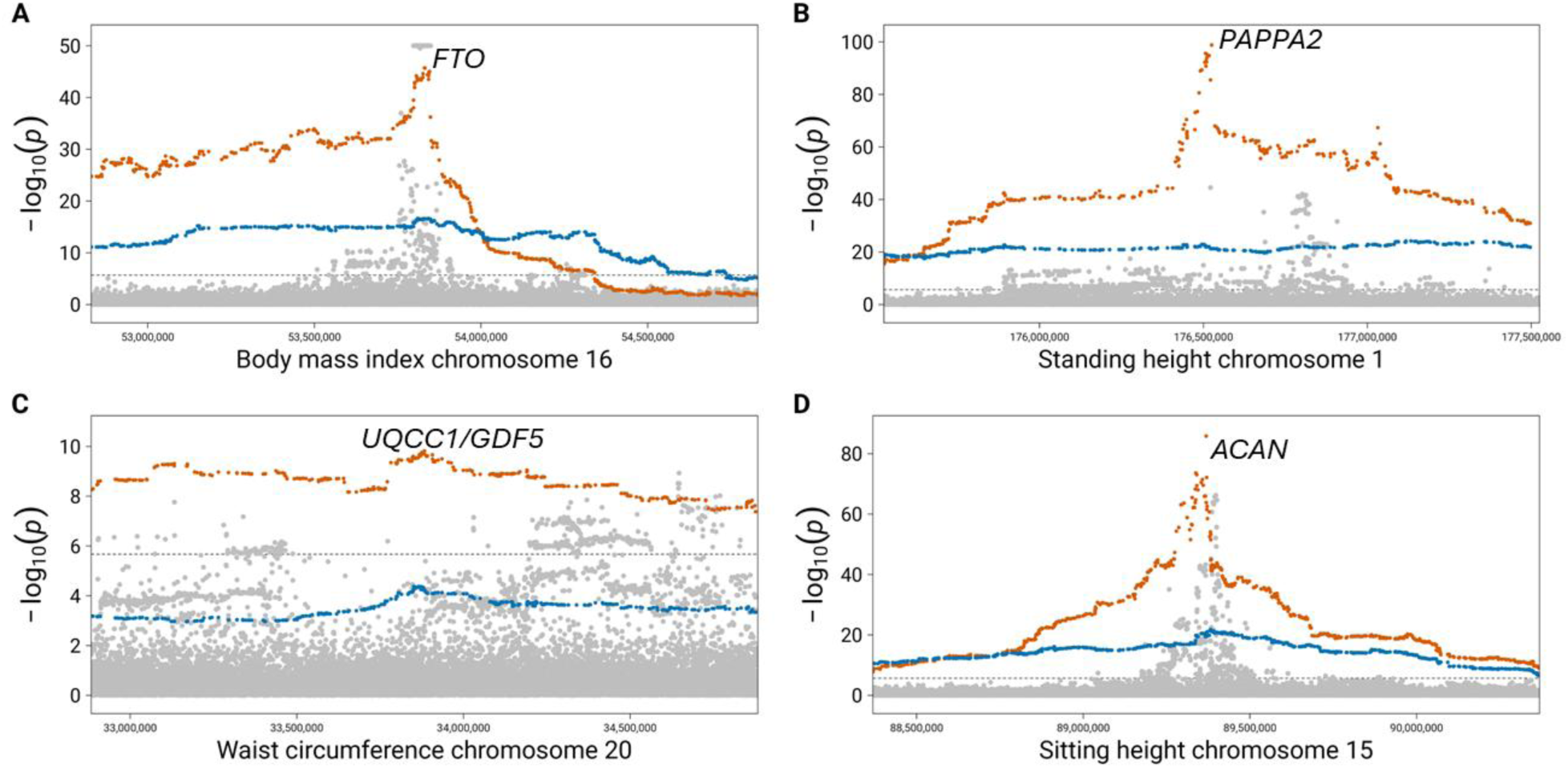
GWAS (grey), HiFiMAP 3 cM (blue), and HiFiMAP 2 cM (orange) regional association plots. (A) Body mass index at the *FTO* locus on chromosome 16, GWAS p-values were truncated at 10^−50^; (B) Standing height at the *PAPPA2* locus on chromosome 1; (C) Waist circumference at the *UQCC1*/*GDF5* locus on chromosome 20; and (D) Sitting height at the *ACAN* locus on chromosome 15. The dashed line represents the significance threshold for HiFiMAP (2 cM UKB white British 410k^34^): 2.13 × 10^−6^.

To contextualize these regional findings globally, we summarized the total number of genome-wide significant loci across the six anthropometric traits (Table 1). While the 2 cM threshold provided amplified power at specific established loci (Fig 5), the loci counts reveal a highly trait-dependent genetic architecture. For traits such as hip circumference, body weight and BMI, the 2 cM analysis yielded a higher number of significant loci. Conversely, for highly polygenic traits like standing height and sitting height, the 3 cM analysis identified a higher number of significant loci. Furthermore, we assessed the number of significant IBD mapping loci that lacked GWAS evidence. We took a conservative approach by excluding IBD mapping loci that were located within 2 cM or 3 cM of a significant GWAS locus from the 2 cM and 3 cM IBD mapping analyses, respectively. After excluding these loci, the 3 cM analysis consistently exhibited a higher number of loci for standing height and sitting height, while the 2cM analysis results had a higher number of loci for hip circumference and body weight.

**Table 1.** Numbers of significant loci in UKB High-resolution IBD mapping (2 cM and 3 cM) and GWAS for 6 anthropometric traits. Genomic loci for all three analyses were defined by grouping contiguous significant variants within 250 kb of one another. IBD mapping loci not within 2 cM or 3 cM flanking regions of any GWAS loci were classified as “No GWAS evidence”.

| Trait | 2cM | 3cM | 2 cM<br>(No GWAS<br>evidence) | 3 cM<br>(No GWAS<br>evidence) | GWAS |
| --- | --- | --- | --- | --- | --- |
| Waist<br>circumference | 53 | 60 | 5 | 11 | 475 |
| Hip<br>circumference | 55 | 47 | 7 | 4 | 507 |
| Standing height | 421 | 452 | 29 | 52 | 1016 |
| Sitting height | 238 | 277 | 6 | 10 | 849 |
| Body mass index | 100 | 86 | 9 | 10 | 629 |
| Body weight | 68 | 49 | 3 | 1 | 690 |

### Application to high-dimensional UDIPs

To demonstrate HiFiMAP’s utility in analyzing high-dimensional brain imaging data, we applied it to the T1 (N = 36,241) and T2 (N = 35,111) 128-dimensional UDIPs. For this genome-wide analysis, we utilized IBD segments with a minimum length of 1 cM to maximize the detection of shorter, ancestral shared haplotypes, and aggregated the association signals across the 128 phenotypes using a minP approach. Figure 6 illustrates the resulting Manhattan plots, revealing multiple genome-wide significant loci across both structural modalities. The T1 analysis (Fig 6A) identified signals mapping to *NR2F1* on chromosome 5 (associated with brain region volumes and shape^6,40^), *SCGN/HIST1H/SLC17A4* on Chromosome 6 (implicated in white matter microstructure and schizophrenia^41,42^), *LINC01163* on Chromosome 10 (associated with Alzheimer’s disease (AD) and total PHF-tau^43,44^), *NUAK1* on Chromosome 12 (associated with attention deficit hyperactive disorder (ADHD)^45^), and *NSF/WNT3* on chromosome 17 (associated with Parkinson’s disease (PD) risk and general cognitive ability^46,47^). Similarly, the T2 analysis (Fig 6B) detected significant associations at *TAL1* on chromosome 1 (associated with major depressive disorder and neuroticism^48,49^) and *CCDC91* on chromosome 10, a gene highly expressed in the central nervous system whose variations and protein interactions are implicated in bipolar disorder^50^, AD pathophysiology^51,52^, and treatment-resistant schizophrenia^50^. Also, there were shared signals identified at the *SCGN* and *NUAK1* loci for T2.

**Fig 6.**
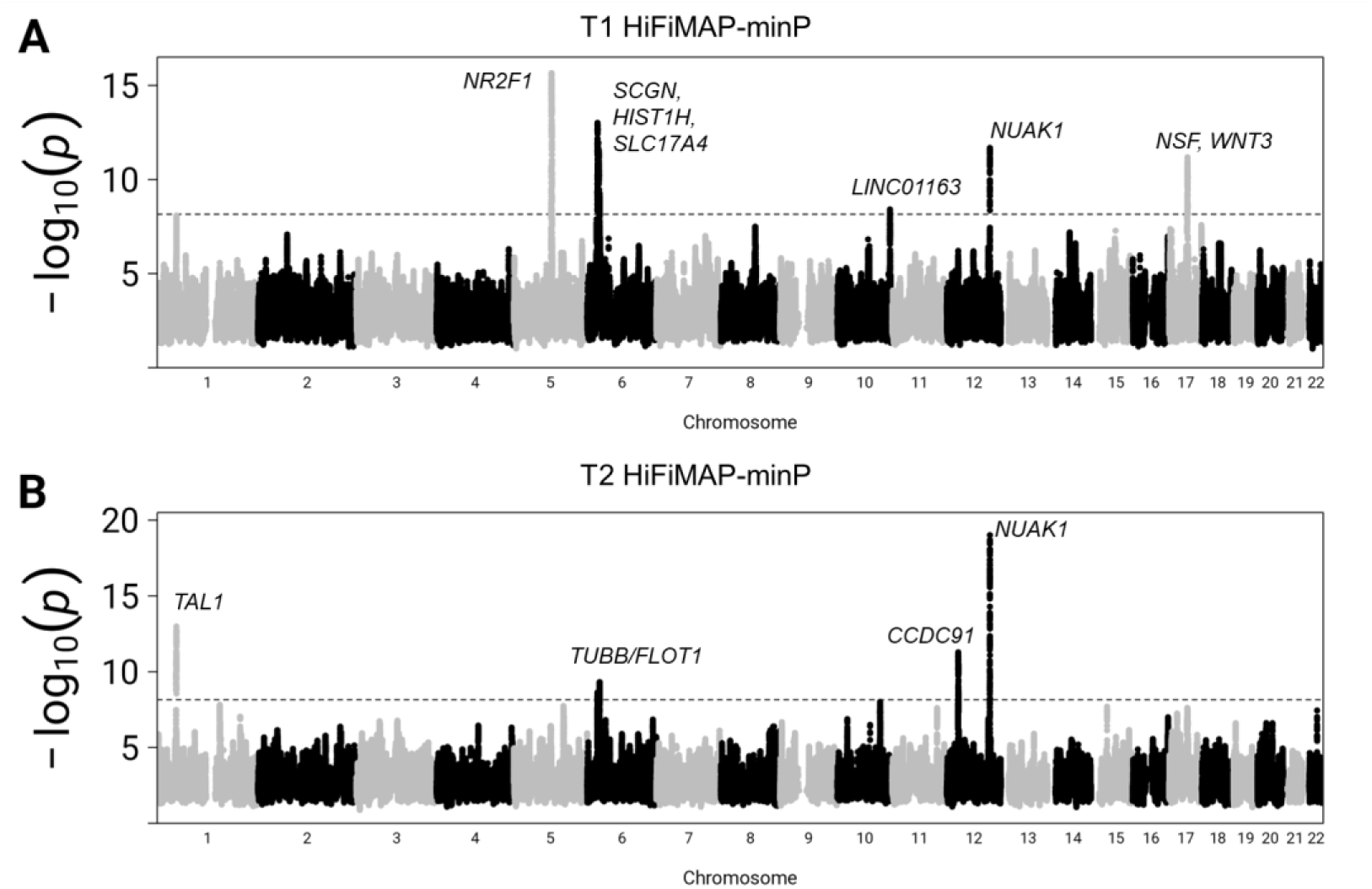
Manhattan plots of the T1 and T2 HiFiMAP analyses applied to 128 high-dimensional UDIPs. Results were generated using a minP approach across 1 cM IBD segments. (A) The T1 HiFiMAP-minP results. (B) The T2 HiFiMAP-minP results. The dashed line represents the Bonferroni corrected genome-wide significance threshold for HiFiAMP 1 cM (N = 36k) using the simulation approach^34^: 8.93 × 10^−7^/256 = 3.49 × 10^−9^.

To further investigate the capabilities of this approach, we compared the regional association signals from the T1 HiFiMAP analysis against those obtained from a conventional single-variant GWAS (GWAS-minP). The regional association plots for the four identified T1 loci are shown in Figure 7. Across the *NR2F1* locus (Fig 7A), the *SCGN/SLC17A4* region (Fig 7B), the *NUAK1* locus (Fig 7C), and the *NSF/WNT3* locus (Fig 7D), the HiFiMAP-minP signals clearly surpass the 1 cM Bonferroni-corrected genome-wide significance threshold (3.49 × 10^−9^). In contrast, the corresponding GWAS-minP results at *NR2F1*, *SCGN/SLC17A4*, *and NSF/WNT3* are not significant at the Bonferroni corrected GWAS significance level for testing 256 phenotypes at 5 × 10^−8^/256 = 1.95 × 10^−10^.

**Fig 7.**
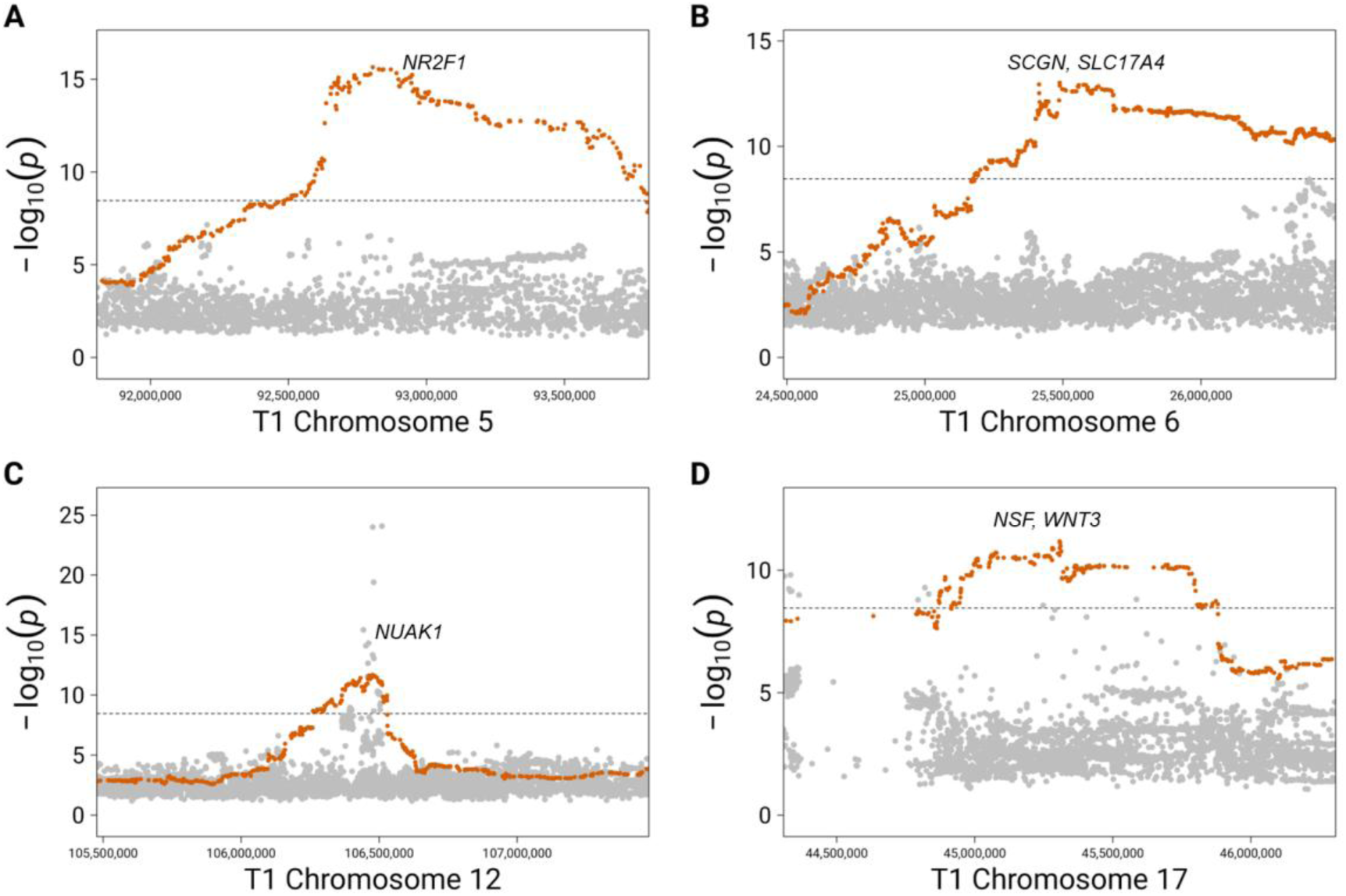
GWAS-minP (grey) and HiFiMAP-minP (orange) regional association plots for T1. (A) Chromosome 5 (*NR2F1*), (B) Chromosome 6 (*SCGN, SLC17A4*), (C) Chromosome 12 (*NUAK1*), and (D) Chromosome 17 (*NSF, WNT3*). The dashed line represents the Bonferroni corrected genome-wide significance threshold for HiFiMAP 1 cM (N = 36k) using the simulation approach^34^: 8.93 × 10^−7^/256 = 3.49 × 10^−9^.

A comprehensive summary of all genome-wide significant loci, including the specific UDIP dimensions driving each signal and their comparative baseline metrics is provided in Table 2. Crucially, Table 2 also reports the HiFiMAP association strength after conditioning on the lead GWAS variant at each locus. Conditioning on the lead GWAS variant was only performed if a suggestive genome-wide significant GWAS SNP was actually present in the ± 1 megabase (Mb) region of each HiFiMAP peak (defined by an unadjusted threshold of 5 × 10^−8^). For the vast majority of identified loci, particularly across the T1 analysis, the HiFiMAP signals remain significant after conditioning (accounting for multiple testing, Bonferroni-corrected threshold of 0.05/9). This indicates that the IBD mapping approach is capturing novel, independent haplotypic architectures or rare-variant clusters that are distinct from the top marginal GWAS signals. The notable exception is the *NUAK1* locus in the T2 analysis, where the conditional p-value attenuates to non-significance (0.34) suggesting the IBD signal at this specific locus is largely driven by the known common variant. Overall, these comparisons underscore the unique strength of HiFiMAP to uncover complementary genetic association findings that influence brain morphology but are systematically missed by standard single-variant GWAS approaches.

**Table 2.** Significant genomic loci associated with T1 and T2 UDIPs identified via HiFiMAP-minP. Summary of genome-wide significant findings from the 1 cM HiFiMAP analysis of 128 T1/T2 UDIPs. GWAS-minP and conditional HiFiMAP p-values are included to assess the independence of the IBD-based regional signals from the GWAS variants. *These loci had no GWAS p-values < 5 × 10^−8^ in the 2 Mb flanking region; conditioning was not performed

| Chr | Pos | P-value<br>(HiFiMAP-minP) | UDIPs<br>dimension | Mapped<br>genes | Neurological<br>disorders/traits | P-value<br>(GWAS-minP) | P-value<br>(conditional) |
| --- | --- | --- | --- | --- | --- | --- | --- |
| 5 | 92805573 | $2.34 \times 10^{-16}$ | T1 30 | NR2F1* | Brain region<br>volumes, brain<br>shape | $2.79 \times 10^{-7}$ | $2.34 \times 10^{-16}$ |
| 6 | 25488265 | $9.67 \times 10^{-14}$ | T1 89 | SCGN/<br>HIST1H/<br>SLC17A4 | White matter<br>microstructure,<br>schizophrenia | $3.65 \times 10^{-9}$ | $1.33 \times 10^{-2}$ |
| 10 | 130010650 | $3.47 \times 10^{-9}$ | T1 68 | LINC01163* | Alzheimer's<br>disease, total<br>PHF-tau | $4.04 \times 10^{-6}$ | $3.47 \times 10^{-9}$ |
| 12 | 106477376 | $2.09 \times 10^{-12}$ | T1 110 | NUAK1 | Attention deficit<br>hyperactive<br>disorder | $8.37 \times 10^{-25}$ | $5.57 \times 10^{-3}$ |
| 17 | 45307359 | $6.60 \times 10^{-12}$ | T1 59 | NSF/WNT3 | Parkinson's<br>disease, general<br>cognitive ability | $5.19 \times 10^{-10}$ | $5.82 \times 10^{-2}$ |
| 1 | 47873311 | $1.03 \times 10^{-13}$ | T2 121 | TAL1 | Major depressive<br>disorder,<br>neuroticism | $4.73 \times 10^{-25}$ | $2.20 \times 10^{-3}$ |
| 6 | 30766945 | $4.83 \times 10^{-10}$ | T2 48 | TUBB/FLOT1* | Schizophrenia,<br>general cognitive<br>ability | $4.91 \times 10^{-6}$ | $4.83 \times 10^{-10}$ |
| 12 | 28235066 | $5.18 \times 10^{-12}$ | T2 114 | CCDC91 | Bipolar disorder,<br>AD<br>pathophysiology,<br>treatment-resistant<br>schizophrenia | $9.09 \times 10^{-37}$ | $2.51 \times 10^{-6}$ |
| 12 | 106511173 | $9.80 \times 10^{-20}$ | T2 42 | NUAK1 | Attention deficit<br>hyperactive<br>disorder | $4.68 \times 10^{-29}$ | $3.35 \times 10^{-1}$ |

### Computational efficiency benchmark

To evaluate the computational efficiency of HiFiMAP, we benchmarked its performance against the original window-based FiMAP method using the full-scale UKB anthropometric cohort (N = 407,681). Despite testing at a vastly higher resolution compared to FiMAP’s 3,403 1 cM windows, HiFiMAP demonstrated extraordinary computational speed. For the 3 cM analysis in the massive UKB anthropometric cohort, testing 640,899 SNPs required only 344 CPU hours, averaging 1.92 seconds per test. In contrast, while FiMAP completed its analyses in 90 CPU hours, its per-test computational burden was drastically higher (95.2 seconds per 1 cM window).

## Discussion

In this study, we introduced the HiFiMAP, a novel variance component-based approach that simultaneously achieves single-SNP resolution and optimized computational efficiency for IBD mapping. Existing IBD mapping frameworks, such as FiMAP and a recent likelihood ratio test developed by Cai and Browning^7,24^, have been instrumental in capturing the effects of untyped rare variants and long-range haplotypes that are frequently missed by standard single-variant GWAS. However, their reliance on fixed genomic windows (e.g., 1 cM chunks) has limited their mapping resolution, hindering precise loci identification and subsequent biological interpretation. HiFiMAP effectively overcomes these limitations, offering a scalable, high-resolution solution for modern biobank-scale datasets. A primary achievement of HiFiMAP is its extraordinary computational efficiency. The exponential increase in the density of the IBD sharing matrix especially when utilizing shorter IBD segments like 2 cM for biobank scale datasets pushes traditional window-based methods to their computational limits. By implementing a dynamic updating algorithm (Fig 1), HiFiMAP efficiently modifies the sparse IBD matrix between adjacent SNPs by only adding or removing segments that start or end at the current position, rather than recalculating the entire matrix from scratch. Coupled with a program architecture optimized for parallelized chunk processing and local IBD matrix checkpoints, this approach drastically reduces redundant memory allocation and increases computing efficiency. This is evidenced by our benchmark in the UKB anthropometric cohort (N > 400,000), where HiFiMAP averaged just 1.92 seconds per test, a striking improvement over the 95.2 seconds per test required by the original FiMAP. Consequently, HiFiMAP enables researchers to conduct true genome-wide IBD mapping at SNP-level resolution without prohibitive computational bottlenecks.

Beyond its computational efficiency, our simulations confirm that the method maintains well-calibrated type-I error rates. While a mild deflation was observed at the extreme tail of the distribution (Fig 2); this is a recognized phenomenon in IBD mapping^24,34,39^, particularly in smaller cohorts (N = 5000). Because IBD segments span continuous genomic regions, the test statistics are highly correlated across adjacent loci, causing the resulting p-values to decay slowly and preventing the independent, random spikes in significance typically seen in single-variant GWAS. Furthermore, the inherent sparsity of local IBD sharing in small cohorts makes it difficult to produce large numbers of extreme p-values on the tail. Together, this spatial correlation and sparse sharing restrict the ability to generate extreme tail p-values under the null hypothesis, visually manifesting as a deflated tail. Power simulation confirmed that HiFiMAP has superior statistical power in detecting rare-variants and haplotype effects compared to the standard GWAS approach, especially when including shorter IBD segments (1 cM HiFiMAP).

Our empirical analyses of UKB anthropometric traits demonstrated the enhanced statistical power of lowering the minimum IBD segment length threshold for well-established loci. While our 3 cM HiFiMAP analysis successfully replicated known FiMAP baselines, transitioning to a 2 cM threshold not only uncovered novel association signals, but also yielded stronger signals for established loci compared to the 3 cM analysis and traditional single-variant GWAS (Fig 5). For example, at the *FTO* locus (Supplementary Fig 1) for waist circumference (a well-documented obesity-susceptibility gene^53^), and the *ACAN* locus for sitting height (an essential cartilage structural and anthropometric gene^54–56^), the 2 cM analysis produced much stronger signals than the 3 cM analysis. A similar pattern applied to the *PAPPA2* locus (a key regulator of human growth^57,58^) for standing height, for which the 2 cM HiFiMAP captured significantly stronger associations than standard GWAS. Most interestingly, at the *UQCC1/GDF5* locus for waist circumference, a region heavily implicated in skeletal development and anthropometry^56,59,60^, the 2 cM HiFiMAP results were highly significant, whereas the 3 cM analysis yielded non-significant results and the GWAS produced borderline significance (due to the more stringent significance level). The amplified statistical significance observed in the 2 cM analysis for these well-established loci suggests that the biology underlying these traits involves older, shorter shared genomic blocks. Our global loci counts revealed that highly polygenic traits like height identified more IBD mapping loci from 3 cM analysis to capture massive backgrounds of relatively recent mutations^61,62^. Moving toward IBD mapping is critical for capturing additional insights into haplotypic architectures that standard GWAS approaches are underpowered to detect.

Building on this new tool, we pushed the mapping resolution even further by applying a 1 cM HiFiMAP threshold to the 128-dimensional T1 and T2 UDIPs. Aggregating signals via a minP approach, this 1 cM analysis successfully uncovered 5/4 genome-wide significant loci across T1/T2 UDIPs, and 4/1 of them were missed by standard GWAS. The T1 analysis identified prominent signals mapping to *NR2F1*, the *SCGN/HIST1H/SLC17A4* cluster, *LINC01163*, *NUAK1*, and *NSF/WNT3*, genes deeply implicated in severe neurological phenotypes ranging from schizophrenia and PD to ADHD and AD pathophysiology^6,40–45^. Similarly, the T2 analysis detected additional significant associations for loci highly involved in schizophrenia and the central nervous system: *TUBB/FLOT1*, *TAL1* and *CCDC91*^42,48–52,63^. Importantly, conditioning on the lead GWAS variants at these loci revealed that 3 out of 4 of our HiFiMAP signals remained significant. This conditional independence proves that our IBD mapping approach is not merely redundantly tagging common variants, but is successfully capturing novel, unmeasured haplotypic architectures or rare-variant clusters influencing brain morphology that are missed by traditional single-variant models.

While our 1 cM HiFiMAP-minP analysis successfully illuminated these highly relevant neurobiological loci, the minP approach itself remains inherently univariate. It tests each UDIP dimension separately, thus suffering from a stringent Bonferroni correction for multiple testing across 128 phenotypes. Because neuroimaging phenotypes are highly polygenic and exhibit complex, correlated genetic architectures, applying a univariate method combined with strict multiple-testing penalties is fundamentally conservative and may leave weaker pleiotropic signals undiscovered. To fully capture these pleiotropic effects and further maximize statistical power across such dense, high-dimensional datasets, a future direction will be the development of a scalable multivariate extension to our framework capable of jointly modeling phenotypic covariance.

Nevertheless, the foundational advancement of the current HiFiMAP framework remains its extraordinary computational scalability and efficiency. By overcoming the severe memory and runtime bottlenecks of traditional window-based methods, HiFiMAP makes SNP-level IBD mapping at 2 cM threshold a practical reality for massive, biobank-scale cohorts (N > 400,000). As highlighted by our enriched anthropometric and neuroimaging discoveries, this optimized, ultra-efficient framework significantly enhances both the discovery yield and the fine-mapping resolution of complex traits, providing a powerful and highly efficient tool for dissecting the rare variant and extended haplotypic architectures of human disease.

## Data availability

The GWAS summary statistics used in this study have been uploaded to the GWAS Catalog https://www.ebi.ac.uk/gwas/ with study accession ID GCST90455631, GCST90455632, GCST90455633, and GCST90455634. The UDIPs are being uploaded to http://deependo.org. UKB data was accessed via approved project 24247.

## Code availability

The code is available via https://github.com/baihongguo/HiFiMAP (GPLv3 license).

## Declaration of interests

HC received consulting fees from Character Biosciences.

## Acknowledgments

This work was supported by grants from the National Institute on Aging (U01 AG070112 and R01 AG081398).

## Notes

### Competing Interest Statement

The authors have declared no competing interest.

### Author Declarations

Committee for the protection of human subjects of The University of Texas Health Science Center at Houston gave ethical approval for this work under No. HSC-SBMI-20-1323 and HSC-SPH-23-0685.

