## Supplementary Information for "HiFiMAP: High-resolution fast identity-by-descent mapping test"

### 1. Supplementary Figures 2. Supplementary Table

### Supplementary Figures

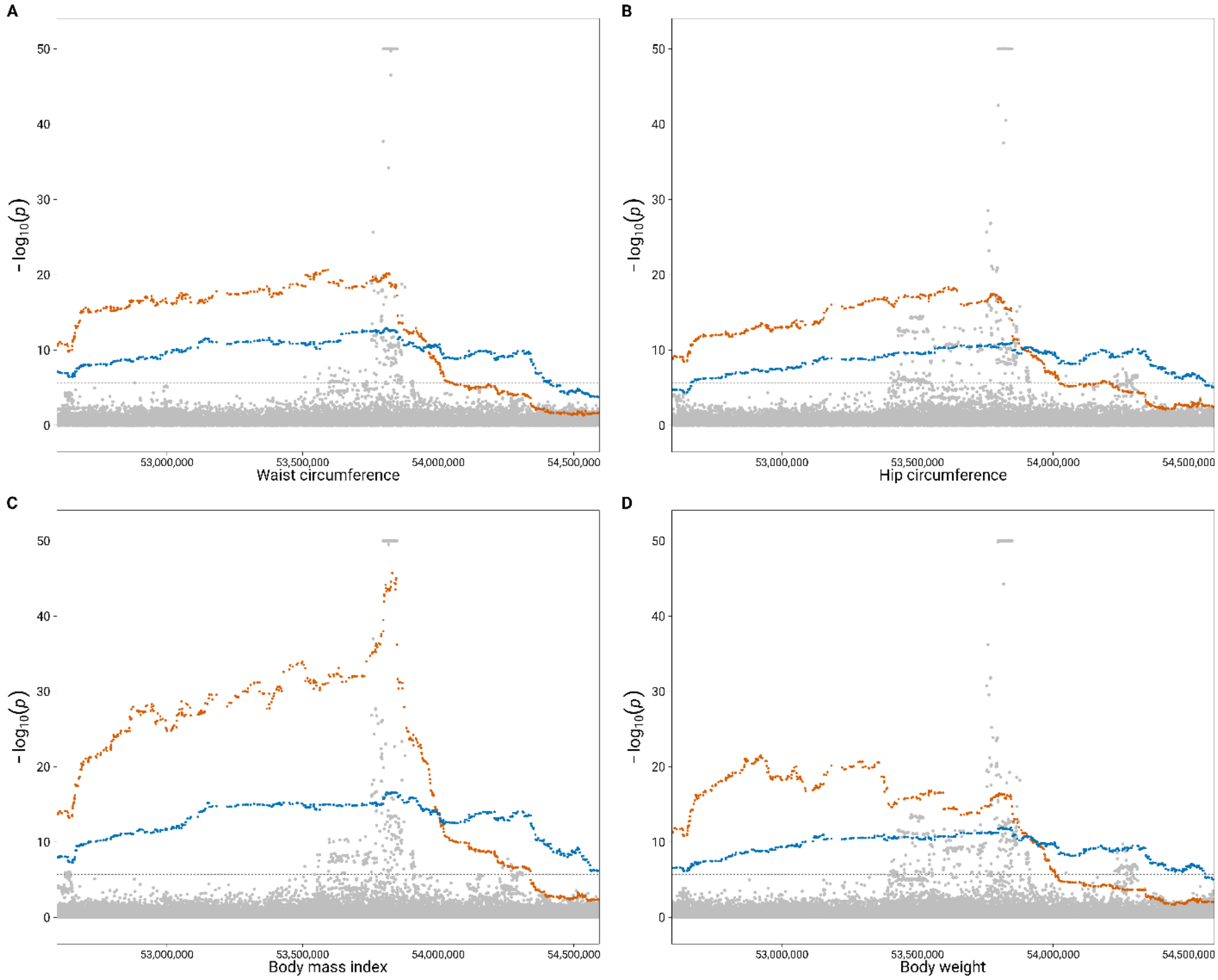


**Supplementary Fig 1. Regional manhattan plots of 2cM (blue), 3cM (orange) HiFiMAP and GWAS (grey) results of 4 anthropometric traits for *FTO* region on chromosome 16**. The dashed line represents the significance threshold of $2.13 \times 10^{-6}$ for 2cM HiFiMAP analysis. GWAS p-values were truncated at $10^{-50}$.

### Supplementary Tables

| Length threshold | Estimated Decay parameter | Threshold (Browning's simulation approach) | Threshold (Empirical null simulation, 200 replicates) |
|---|---|---|---|
| 1cM | 35.5 | $2.32 \times 10^{-6}$ | $1.88 \times 10^{-6}$ |
| 2cM | 24.7 | $3.54 \times 10^{-6}$ | $2.87 \times 10^{-6}$ |
| 3cM | 19.3 | $5.27 \times 10^{-6}$ | $4.90 \times 10^{-6}$ |

**Supplementary Table 1. Genome-wide significance thresholds for varying IBD segment lengths.** Estimated decay parameters and the resulting significance cutoffs for HiFiMAP analysis at 1 cM, 2 cM, and 3 cM thresholds. Thresholds were calculated utilizing both Browning's established simulation approach and an empirical null distribution (200 replicates).

| Length threshold | Accuracy | Length accuracy |
|---|---|---|
| 1cM | 0.7627 | 0.7243 |
| 2cM | 0.9994 | 0.9615 |
| 3cM | 0.9999 | 0.9740 |

**Supplementary Table 2. Accuracy of IBD segment in a simulated cohort (N = 5,000).** Accuracy denotes the proportion of true simulated IBD segments correctly identified by the calling algorithm. Length accuracy reflects the precision of the estimated segment boundaries compared to the simulated ground truth.
